# Digital Health Adoption, eHealth Literacy, and Trust in AI Among Generation Z University Students in Sri Lanka: An Empirical Study

**DOI:** 10.64898/2026.07.22.26358733

**Authors:** Susith Athukorala

## Abstract

**Background:** Digital health technologies—spanning mobile applications, telemedicine, and AI-driven platforms—are rapidly reshaping healthcare delivery globally. Although Generation Z university students are classified as digital natives, empirical data evaluating their eHealth literacy, technology acceptance, and specific trust barriers in developing South Asian nations like Sri Lanka remain scarce.

**Objective:** This study evaluated eHealth literacy, technology acceptance, online health information-seeking behaviors, and adoption barriers among Gen Z undergraduates in Sri Lanka, focusing on the interplay between eHealth literacy, AI trust, and digital care preferences.

**Methods:** A cross-sectional survey (N = 172) was conducted among Sri Lankan university undergraduates utilizing adapted, validated instruments: the eHealth Literacy Scale (eHEALS) and the Technology Acceptance Model (TAM). Statistical analysis included scale reliability validation (Cronbach’s alpha), descriptive profiling, Chi-Square (χ^2^) contingency tests, Pearson correlations, and Multiple Linear OLS Regression models.

**Results:** Participants demonstrated high overall eHealth literacy (Mean = 3.84 ± 0.58) and strong endorsement of digital health utility (Mean = 3.99 ± 0.59). Online health searches were reported by 86.6% of respondents. AI tools (e.g., ChatGPT, Gemini) emerged as the second most frequent source for health queries (57.6%), surpassing YouTube (44.2%) and social media (26.2%), with medical students showing significantly higher AI utilization (χ^2^ = 8.70, p = .003). In multiple regression analysis, digital platform preference over physical clinic visits (R^2^ = .352, p < .001) was significantly predicted by Perceived Ease of Use (β = 0.371, p = .001) and Trust in AI Recommendations (β = 0.370, p < .001), whereas face-to-face consultation preference (76.7%) and personal data privacy risks (50.0%) remained predominant adoption barriers.

**Conclusion:** Gen Z students in Sri Lanka exhibit high digital health readiness and substantial reliance on AI-driven information seeking. However, institutional deployment must address privacy concerns and integrate hybrid clinical workflows to bridge the gap between high perceived utility and physical consultation preferences.

**Author Summary:** *Why was this study done?:* Generation Z university students are often called “digital natives,” but we know very little about how young adults in lower-middle-income countries like Sri Lanka actually use digital health apps, online platforms, and AI tools for their personal health.

*What did the researchers do and find?:* We surveyed 172 university students across Sri Lanka using standardized measures of digital health literacy and technology acceptance. We found that 86.6% search for health information online. Surprisingly, conversational AI tools (such as ChatGPT and Gemini) have become the second most popular source for health queries (57.6%), surpassing YouTube, social media, and official government health websites. While students recognize the potential of digital health tools, 76.7% still prefer seeing a doctor face-to-face, primarily due to concerns about personal data privacy and a lack of awareness about local digital services.

*What do these findings mean?:* Young adults are eager to use digital health tools and interactive AI, but high technology access alone does not translate to full adoption of online medical care. To build trust, healthcare systems in Sri Lanka must create easy-to-use, privacy-protected platforms that combine automated digital convenience with professional medical oversight.

## Introduction

The global landscape of healthcare delivery is undergoing a profound digital transformation driven by mobile health (mHealth) applications, wearable monitoring tools, telemedicine platforms, and generative Artificial Intelligence (AI) algorithms [1,2]. In lower-middle-income countries (LMICs), digital health solutions present unprecedented opportunities to democratize healthcare access, reduce systemic costs, and mitigate regional provider shortages [3,4]. Generation Z individuals born roughly between 1995 and 2010 represent the first demographic cohort to mature entirely within a hyper-connected digital ecosystem [5]. Recognized as digital natives, university students within this generation possess high baseline computing competencies, making them early adopters and key stakeholders in digital health ecosystems [5,6].

In Sri Lanka, national survey data indicate high digital literacy rates among young adults, supported by expanding mobile broadband access and widespread smartphone usage [6,7]. During the COVID-19 pandemic, Sri Lankan healthcare systems experienced accelerated digitization, marked by the expansion of national teleconsultation channels (e.g., *e-Channelling, Doc990*) and remote triage services [8]. However, despite this infrastructure evolution, the degree to which Gen Z university students systematically adopt digital health technologies for self-care and medical decision-making remains underexplored.

To evaluate technology adoption dynamics in healthcare, two theoretical models are widely applied: the **Technology Acceptance Model (TAM)** and the **eHealth Literacy Framework** [9,10]. TAM posits that an individual’s intention to use a technology is primarily governed by two cognitive constructs: **Perceived Usefulness (PU)**, the degree to which a user believes a system will enhance health outcomes, and **Perceived Ease of Use (PEOU)**, the extent to which the tool is perceived as effortless [9,11]. Concurrently, eHealth literacy, measured via the **eHEALS** scale, evaluates an individual’s self-efficacy in locating, evaluating, and applying electronic health information to address health issues [10]. High eHealth literacy is hypothesized to alleviate safety concerns and promote active engagement with digital platforms [5].

Despite the theoretical readiness of Gen Z, digital health adoption is hindered by significant systemic and psychological barriers. Globally, youth report persistent concerns regarding data privacy, cybersecurity breaches, misinformation on digital platforms, and skepticism toward automated diagnostic algorithms [12–14]. In developing South Asian contexts, cultural preferences for traditional, face-to-face physician interactions and a lack of local language (Sinhala/Tamil) application interfaces further complicate the digital health adoption landscape [4,7,15]. Furthermore, while conversational AI tools (e.g., ChatGPT, Gemini) have gained rapid traction among students, empirical data regarding how AI trust influences broader digital healthcare acceptance remain lacking.

### Research Objectives

To address these empirical gaps, this study examines digital health engagement among Gen Z university undergraduates in Sri Lanka. Specifically, the study aims to:

1. Assess the baseline **eHealth Literacy (eHEALS)** and **Technology Acceptance (TAM)** metrics among Sri Lankan undergraduates across different fields of study.
2. Identify primary online health information-seeking channels, contrasting traditional web searches with emergent **AI-driven tools**.
3. Evaluate the key predictors (eHEALS, TAM constructs, AI trust, privacy perceptions) that influence a student’s preference for digital health platforms over physical consultations.
4. Quantify the primary perceived barriers and enablers governing digital health utilization in Sri Lanka.

## Methodology

### Ethics Statement

Participation was entirely voluntary, anonymous, and uncompensated. Electronic informed consent was mandatory prior to accessing survey questions. No personally identifiable information (PII), such as names, student IDs, IP addresses, or contact numbers, was collected. The study protocol adhered to the ethical principles of the Declaration of Helsinki and received institutional review board (IRB) exemption.

### Study Design and Setting

A quantitative, cross-sectional online survey was conducted between March 2025 and March 2026 to evaluate digital health adoption, eHealth literacy, technology acceptance, and information-seeking behaviors among Generation Z university students in Sri Lanka. The target population comprised undergraduate students aged 18 to 26 years enrolled in Sri Lankan higher education institutions.

### Sampling and Sample Size

A convenience and snowball sampling methodology was utilized via university email networks, student groups, and digital communication channels (e.g., WhatsApp, Telegram) across public and private university networks. A total of 172 complete, valid responses were recorded (*N* = 172). Based on statistical power calculations for multiple linear regression models with five predictors, a sample size of *N* = 172 exceeds the minimum requirement (*α* = .05, power = .80, medium effect size *f*^2^ = .15), providing sufficient statistical power for parametric testing.

### Survey Instrument and Measurement Scales

The survey instrument was adapted from validated global instruments and structured into six distinct sections:

1. **Demographic Profile:** Captured age, gender, university sector (public vs. private), academic field of study, study year, and self-rated English language proficiency.
2. **eHealth Literacy Scale (eHEALS):** Adapted from Norman and Skinner (2006) [10], utilizing eight items measured on a 5-point Likert scale (1 = Strongly Disagree to 5 = Strongly Agree) to assess self-efficacy in discovering, evaluating, and applying digital health information.
3. **Technology Acceptance Model (TAM):** Adapted from Davis (1989) and Holden and Karsh (2010) [9,11], assessing Perceived Usefulness (PU; 3 items), Perceived Ease of Use (PEOU; 1 item), Behavioral Intention/Preference for Digital Care Over Physical Visits (1 item), Trust in AI Recommendations (1 item), and Data Privacy/Security Trust (1 item).
4. **Health Information National Trends Survey (HINTS):** Adapted from the National Cancer Institute framework [16] to capture 6-month online search activity, primary information channels (e.g., Google, AI tools, social media), source trustworthiness, and online search concerns.
5. **Barriers and Enablers:** Multi-select items identifying primary impediments to frequent digital health usage and systemic factors that would encourage future engagement.
6. **Qualitative Perspectives:** Open-ended prompts soliciting user opinions on improving Sri Lankan digital health systems and future care replacement.

### Data Processing and Statistical Analysis

Data were exported to Python (pandas, statsmodels, scipy.stats) for statistical processing. Scale items were mapped to numeric values (1 = Strongly Disagree to 5 = Strongly Agree). Scale reliability was evaluated using Cronbach’s alpha (*α*), with values ≥ .70 indicating acceptable internal consistency. Continuous variables were summarized using Means and Standard Deviations (± SD), while categorical variables were reported as frequencies and percentages (*N*,%). Statistical associations were evaluated using independent *t*-tests, Chi-Square (*χ*^*2*^) tests of independence, Pearson correlation matrices, and Ordinary Least Squares (OLS) multiple linear regressions. Statistical significance was set at *p* < .05.

## Results

### Sample Demographics

As shown in Table 1, female respondents comprised 65.1% (*n* = 112) of the cohort, while males accounted for 34.9% (*n* = 60). The majority of participants were aged 18–20 years (41.9%, *n* = 72) or 21–23 years (30.8%, *n* = 53). In terms of academic discipline, Medical/Health Sciences/Nursing students constituted 73.8% (*n* = 127), followed by Engineering/IT (22.1%, *n* = 38). Private university undergraduates represented 94.8% (*n* = 163) of the sample. Most respondents rated their English proficiency as Intermediate (59.3%, *n* = 102) or Fluent (25.6%, *n* = 44).

**Table 1:** Demographic Characteristics of the Sample (N = 172)

| **Demographic Variable** | **Category** | **Frequency (N)** | **Percentage (%)** |
| --- | --- | --- | --- |
| **Gender** | Female | 112 | 65.1% |
|  | Male | 60 | 34.9% |
| **Age Group** | Under 18 | 32 | 18.6% |
|  | 18–20 | 72 | 41.9% |
|  | 21–23 | 53 | 30.8% |
|  | 24–26 | 13 | 7.6% |
|  | 27–29 | 2 | 1.2% |
| **Field of Study** | Medical / Health Sciences / Nursing | 127 | 73.8% |
|  | Engineering / Technology / IT | 38 | 22.1% |
|  | Business / Management | 7 | 4.1% |
| **University Type** | Private University | 163 | 94.8% |
|  | Government University | 9 | 5.2% |
| **Study Year** | 1st Year | 99 | 57.6% |
|  | 2nd Year | 54 | 31.4% |
|  | 3rd Year | 15 | 8.7% |
|  | 4th Year or Above | 4 | 2.3% |
| **English Proficiency** | Basic | 26 | 15.1% |
|  | Intermediate | 102 | 59.3% |
|  | Fluent | 44 | 25.6% |

### Scale Reliability and Descriptive Scale Metrics

Both primary composite scales demonstrated strong internal consistency reliability. The 8-item eHEALS composite scale achieved a Cronbach’s alpha of *α* = 0.863, while the 7-item TAM framework scale achieved *α* = 0.800. The sub-dimension for TAM Perceived Usefulness (3 items) yielded *α* = 0.823.

As detailed in Table 2, overall eHealth literacy was high (Mean = 3.84 ± 0.58). Students reported highest confidence in knowing how to find helpful resources online (Mean = 3.91 ± 0.77) and believing digital health is useful for healthcare needs (Mean = 4.07 ± 0.73). Within TAM constructs, Perceived Usefulness items scored high, particularly regarding access improvement (Mean = 4.07 ± 0.70) and national healthcare quality enhancement (Mean = 4.04 ± 0.72). Conversely, Trust in AI Recommendations scored the lowest among all items (Mean = 3.15 ± 0.89).

**Table 2:** Psychometric Scale Items, Descriptives, and Internal Reliability.

| **Construct / Scale Item** | **Mean (1–5)** | **SD** | **Cronbach's $\alpha$** |
| --- | --- | --- | --- |
| **eHealth Literacy Composite (eHEALS, 8 Items)** | **3.84** | **0.58** | **0.863** |
| eHEALS 1: Know how to find helpful health resources online | 3.91 | 0.77 |  |
| eHEALS 2: Know where to find reliable health info online | 3.72 | 0.74 |  |
| eHEALS 3: Differentiate high vs. low-quality health info | 3.72 | 0.71 |  |
| eHEALS 4: Confident using digital tools for health decisions | 3.79 | 0.77 |  |
| eHEALS 5: Know how to use health info to improve health | 3.85 | 0.75 |  |
| eHEALS 6: Comfortable using Sri Lankan platforms (e.g., e-Channelling) | 3.81 | 0.87 |  |
| eHEALS 7: Comfortable using wearable health devices | 3.81 | 0.74 |  |
| eHEALS 8: Believe digital health is useful for health needs | 4.07 | 0.73 |  |
| **Technology Acceptance Constructs (TAM)** |  |  | **0.800** |
| **TAM Perceived Usefulness (PU Composite, 3 Items)** | **3.99** | **0.59** | **0.823** |
| - Digital health is useful in managing health | 3.87 | 0.70 |  |
| - Digital health will improve Sri Lankan healthcare quality | 4.04 | 0.72 |  |
| - Digital health makes accessing healthcare easier | 4.07 | 0.70 |  |
| TAM Perceived Ease of Use (PEOU): Find health apps easy to use | 3.80 | 0.73 | N/A |
| TAM Preference: Prefer digital platform over physical visit | 3.50 | 0.97 | N/A |
| TAM Trust AI: Trust AI recommendations (chatbots/symptom checkers) | 3.15 | 0.89 | N/A |
| TAM Privacy: Believe digital solutions respect privacy & security | 3.49 | 0.78 | N/A |

**Table 3:** OLS Multiple Linear Regression Models.

| Dependent Variable | Independent Predictor | Unstandardized Coeff ($\beta$) | SE | t-value | p-value | Model Diagnostics |
| --- | --- | --- | --- | --- | --- | --- |
| Model 1:<br>Preference for Digital Platform Over Physical Visit | (Constant) | 0.197 | 0.469 | 0.420 | .675 | $R^2 = .352$ |
| | TAM Perceived Ease of Use (PEOU) | 0.371 | 0.108 | 3.424 | .001* | Adjusted $R^2 = .333$ |
| | TAM Trust in AI Recommendations | 0.370 | 0.081 | 4.549 | <.001* | $F = 18.07, p < .001$ |
|  | TAM Privacy & Security Trust | 0.146 | 0.092 | 1.593 | .113 |  |
|  | eHEALS Score | 0.073 | 0.171 | 0.430 | .668 |  |
|  | TAM Perceived Usefulness (PU) | -0.015 | 0.145 | -0.102 | .919 |  |
| Model 2:<br>Perceived Usefulness (PU) | (Constant) | 0.821 | 0.243 | 3.385 | .001 | $R^2 = .522$ |
| | eHEALS Score | 0.625 | 0.077 | 8.088 | <.001* | Adjusted $R^2 = .511$ |
| | TAM Perceived Ease of Use (PEOU) | 0.187 | 0.056 | 3.337 | .001* | $F = 45.65, p < .001$ |
|  | TAM Trust in AI Recommendations | -0.015 | 0.043 | -0.339 | .735 |  |
|  | TAM Privacy & Security Trust | 0.032 | 0.049 | 0.658 | .512 |  |

\*Significant at $p < .01$ .

### Health Information-Seeking Patterns and AI Adoption

An overwhelming 86.6% (*n* = 149) of respondents searched for health information online in the past 6 months. Regarding source trustworthiness, 55.8% rated online health info as “Somewhat Trustworthy,” 33.1% were “Neutral,” and only 8.1% deemed it “Very Trustworthy”.

As illustrated in Table 4, while **Google/Web Search** was the most frequented source (84.3%, *n* = 145), **Generative AI tools (ChatGPT, Gemini, Copilot)** emerged as the **second most utilized channel (57.6%**, *n* = 99), outranking YouTube/Videos (44.2%), medical websites (27.3%), social media platforms (26.2%), and government health sites (22.1%). Chi-Square analysis revealed that Medical/Health Sciences students were significantly more likely to utilize AI tools for health queries than non-medical students (67.2% vs. 37.8%; *χ*^*2*^ = 8.70,*p* = .003).

**Table 4:** Health Search Sources, Online Concerns, Barriers, and Motivators (N = 172)

| Category | Variable / Item | Frequency (N) | Percentage (%) |
| --- | --- | --- | --- |
| Health Info Sources<br>(Select all that apply) | Google / Web Search | 145 | 84.3% |
|  | AI Tools (ChatGPT, Gemini, Copilot, etc.) | 99 | 57.6% |
|  | YouTube / Online Videos | 76 | 44.2% |
|  | Medical Websites (WebMD, UpToDate, etc.) | 47 | 27.3% |
|  | Social Media (Facebook, Instagram, TikTok, Twitter) | 45 | 26.2% |
|  | Government Health Websites (MOH Sri Lanka) | 38 | 22.1% |
|  | Health Mobile Apps (e-Channelling, Apple Health, etc.) | 36 | 20.9% |
| Online Search Concerns (Max 4 options) | Misinformation or fake news | 121 | 70.3% |
|  | Privacy and security risks | 95 | 55.2% |
|  | Lack of credibility in sources | 76 | 44.2% |
|  | Lack of medical professional oversight | 71 | 41.3% |
|  | Outdated information | 55 | 32.0% |
|  | Difficulty understanding medical jargon | 50 | 29.1% |
|  | Commercial bias and conflicts of interest | 40 | 23.3% |
| Primary Barriers (Max 4 options) | Preference for face-to-face consultations | 132 | 76.7% |
|  | Concerns about personal data security | 86 | 50.0% |
|  | Lack of awareness about available services | 75 | 43.6% |
|  | Poor quality of service / connectivity | 55 | 32.0% |
|  | General distrust of online platforms | 51 | 29.7% |
|  | Limited access to smartphones/computers | 33 | 19.2% |
|  | Internet/data costs are too high | 27 | 15.7% |
| Key Enablers (Max 4 options) | Ease of Use and Accessibility | 90 | 52.3% |
|  | Reliable info & professional endorsements | 87 | 50.6% |

### Predictors of Digital Platform Preference and Perceived Usefulness

Multiple linear regression analysis (OLS) was conducted to identify key drivers of digital technology preference and utility.

In **Model 1** (*R*^2^ = .352,*F*(5,166) = 18.07,*p* < .001), predicting preference for using a digital platform over a physical hospital/clinic visit:

- **Perceived Ease of Use (PEOU)** was a strong, positive, statistically significant predictor (*β* = 0.371,*t* = 3.424,*p* = .001).
- **Trust in AI Recommendations** was also a strong, positive, statistically significant predictor (*β* = 0.370,*t* = 4.549,*p* < .001).
- eHEALS scores (*β* = 0.073,*p* = .668), Perceived Usefulness (*β* = ―0.015,*p* = .919), and Privacy Trust (*β* = 0.146,*p* = .113) were not independent direct predictors of digital consultation preference.

In **Model 2** (*R*^2^ = .522,*F*(4,167) = 45.65,*p* < .001), predicting overall TAM Perceived Usefulness (PU):

- **eHealth Literacy (eHEALS)** was the strongest positive predictor (*β* = 0.625,*t* = 8.088,*p* < .001).
- **Perceived Ease of Use (PEOU)** was also significant (*β* = 0.187,*t* = 3.337,*p* = .001).

### Barriers and Enablers to Digital Health Adoption

The predominant barrier to digital health adoption was a strong entrenched preference for face-to-face doctor consultations (76.7%, *n* = 132). Personal data security concerns (50.0%, *n* = 86) and lack of awareness regarding available local services (43.6%, *n* = 75) were also major impediments.

Regarding adoption enablers, respondents selected **Ease of Use and Accessibility (52.3%**, *n* = 90**), Professional Endorsements and Reliable Content (50.6%**, *n* = 87), and **Affordable/Free Digital Services (45.9%**, *n* = 79) as the top motivators.

## Discussion

This study evaluated digital health adoption, eHealth literacy, technology acceptance constructs, and AI reliance among Generation Z university students in Sri Lanka. The findings provide key empirical insights into how youth in a lower-middle-income country (LMIC) interact with digital health tools. While respondents demonstrated high baseline eHealth literacy (Mean = 3.84) and overwhelmingly endorsed the utility of digital health systems for expanding healthcare access (Mean = 4.07), actual adoption remains constrained by structural privacy fears and a strong preference for face-to-face physician care.

A notable finding from this dataset is the high reliance on conversational Artificial Intelligence (AI) tools (e.g., ChatGPT, Gemini) for health information seeking. Over 57% of students reported using generative AI as a primary channel for health queries, outranking YouTube, medical websites, social media platforms, and government health portals. Medical and health sciences undergraduates exhibited significantly higher AI utilization compared to non-medical peers (*χ*^*2*^ = 8.70,*p* = .003). This rapid integration highlights a shift in how digital natives seek health information, moving away from static web searches toward interactive, conversational synthesis [5,14]. However, this high reliance on AI tools exists alongside low baseline trust in automated diagnostic recommendations (Mean = 3.15) and significant concerns regarding online misinformation (70.3%). Students appear to leverage AI platforms for rapid informational triage while maintaining a skeptical attitude toward automated clinical decision-making [12,13].

In the multiple linear regression models, digital platform preference over physical consultation visits (*R*^2^ = .352) was significantly driven by Perceived Ease of Use (*β* = 0.371,*p* = .001) and Trust in AI Recommendations (*β* = 0.370,*p* < .001). Interestingly, overall eHealth literacy (eHEALS) and Perceived Usefulness (PU) were not direct predictors of digital consultation preference. Instead, eHealth literacy acted as a strong foundational antecedent that strongly predicted Perceived Usefulness (*β* = 0.625,*p* < .001 in Model 2). These findings align with extended Technology Acceptance Model (TAM) literature in healthcare [4,9,11]. They suggest that while eHealth literacy equips students to recognize the value of digital platforms, actual willingness to replace a physical clinic visit depends heavily on system interface simplicity and cognitive trust in the underlying technology.

Despite high technology readiness, 76.7% of respondents reported a persistent preference for face-to-face medical consultations. This preference reflects both cultural norms in South Asia regarding clinical interactions and ongoing concerns over personal data privacy (50.0%) and service quality (32.0%). In Sri Lanka’s healthcare environment, patient-provider relationships are traditionally built on direct physical examination and interpersonal rapport [7,8]. Furthermore, 43.6% of students cited a lack of awareness regarding existing local digital health platforms (such as *e-Channelling* or public hospital portals). This reveals a disconnect between the availability of national digital health infrastructure and youth engagement. Bridging this gap requires transitioning from standalone digital apps toward integrated, hybrid care models that preserve provider rapport while streamlining administrative and routine monitoring tasks.

### Actionable Recommendations for Health Informaticians

Health informaticians, digital health architects, and software developers must prioritize the co-design of user-centered interfaces that emphasize simplicity, intuitive navigation, and low-cognitive load. Because Perceived Ease of Use emerged as a primary driver of digital care preference among Gen Z, applications should avoid complex registration workflows, high bandwidth requirements, or dense medical jargon. System designs should incorporate clear visual dashboards, localized content in Sinhala and Tamil alongside English, and lightweight architectures capable of operating efficiently across variable mobile network conditions in both urban and rural Sri Lankan regions.

To address the major barrier of data privacy concerns, healthcare system architects and policymakers must establish robust, transparent data governance frameworks aligned with the Sri Lanka Personal Data Protection Act (PDPA No. 9 of 2022). Digital health platforms should visibly incorporate end-to-end encryption, explicit opt-in consent protocols, and clear explanations of how personal health data is stored and utilized. Health informaticians should also work closely with medical institutions to secure official endorsements and professional oversight badges on certified health platforms. Demonstrating compliance with national cybersecurity standards will help mitigate user privacy risks and build public trust in digital health platforms.

Given that Gen Z students frequently use AI tools for health queries while maintaining low trust in automated diagnostics, informaticians should focus on deploying hybrid, clinician-in-the-loop AI systems rather than fully autonomous diagnostic tools. Integrating conversational AI assistants into verified patient portals, where AI-generated insights are reviewed or triaged by qualified medical professionals, can combine the speed of conversational search with clinical reliability. Furthermore, educational institutions and health informaticians should collaborate to integrate digital health literacy and AI evaluation modules into university curricula, empowering students to critically evaluate algorithm-driven health advice and navigate national digital health resources effectively.

## Conclusion & Limitations

## Conclusion

Generation Z university students in Sri Lanka demonstrate high eHealth literacy, positive perceptions of digital health utility, and strong engagement with emerging AI-driven health tools. However, widespread adoption of digital care platforms is tempered by data privacy concerns, a lack of awareness regarding local digital services, and a preference for physical clinical consultations. System preference is primarily driven by interface simplicity and trust in AI recommendations. To capitalize on Gen Z’s high digital readiness, healthcare stakeholders in Sri Lanka must develop localized, privacy-focused, hybrid digital health solutions that combine user-friendly design with professional clinical oversight.

### Limitations and Future Research

This study has several limitations that should be acknowledged. First, the sample size of *N* = 172, while statistically sufficient for parametric testing and linear regression models, was predominantly drawn from private university undergraduates and medical/health science disciplines. This may limit the generalizability of the findings to public university cohorts or non-academic youth in rural Sri Lanka. Second, the cross-sectional design captures user attitudes at a single point in time, preventing causal conclusions regarding long-term technology adoption. Third, data on technology adoption were self-reported, which may introduce self-assessment bias. Future research should employ longitudinal designs, expand sampling across public state universities in all provinces, and utilize objective usage metrics (e.g., system log data) to evaluate real-world digital health engagement in Sri Lanka.

### Declaration of Generative AI and AI-Assisted Technologies in the Writing Process

During the preparation of this work, the author utilized generative artificial intelligence technologies to assist in refining technical prose, structuring tabular data outputs, and assisting with statistical code execution in Python. After using these services, the author thoroughly reviewed, edited, and verified all statistical outputs, tables, and narrative text, and takes full responsibility for the final scientific content and accuracy of the manuscript.

## Acknowledgments

The author wishes to acknowledge the university students across Sri Lanka who voluntarily participated in this study, as well as the open-source digital health community for providing validated research frameworks (eHEALS and TAM) that made this empirical evaluation possible.

## Data Availability Statement

The anonymized quantitative dataset (*N* = 172) and statistical analysis scripts supporting the findings of this study are provided in **S1 Data**.

## Supporting Information

S1 Data. Anonymized Survey Dataset (N = 172). Excel spreadsheet (.xlsx) containing raw respondent data, eHEALS scores, TAM scales, and information-seeking variables analyzed in this study.

